# A systematic review of the concept “male involvement in maternal health” by natural language processing and descriptive analysis

**DOI:** 10.1101/2021.01.13.21249728

**Authors:** Anna Galle, Gaëlle Plaieser, Tessa Van Steenstraeten, Sally Griffin, Nafissa Osman, Kristien Roelens, Olivier Degomme

## Abstract

**Introduction:** Experts agree that male involvement (MI) in maternal health (MH) is a multifaceted concept, but a universal definition is lacking, hampering comparison of findings and interpretation of the literature. This systematic review aims to examine the conceptualization of MI in MH globally and critically review commonly used indicators.

**Methods:** PubMed, Embase, Scopus, Web of Science and CINAHL databases were searched for quantitative literature (between the years 2000 and 2020) containing indicators or variables representing MI in MH, which was defined as the involvement, participation, engagement or support of men in all activities related to maternal health.

**Results:** After full text review, 282 studies were included in the review. Most studies were conducted in Africa (43%), followed by North America (23%), Asia (15%) and Europe (12%). Descriptive analysis and text mining analysis showed MI in MH has been conceptualised by focusing on two main aspects: psychosocial support and instrumental support for maternal health care utilisation. Differences in measurement and topics were noted according to continent with Africa focusing on HIV prevention, North America and Europe on psychosocial health and stress, and Asia on nutrition. One third of studies used one single indicator for assessing MI in MH and no common pattern of indicators could be identified. Antenatal care attendance was the most used indicator (40%) followed by financial support (17%), presence during childbirth (17%) and HIV testing (14%). Majority of studies did not collect data from men directly.

**Discussion:** Researchers often focus on a single aspect of MI in MH, resulting in the usage of a narrow and simplified set of indicators. Aspects such as communication between the couple, shared decision making, participation in household tasks and the subjective feeling of being supported have received little attention. We believe a more multidimensional approach can broaden the potential of MI programs. Further research, involving experts and pilot testing, is recommended to develop consensus regarding a more robust and comprehensive set of valid and feasible indicators for assessing MI in MH.

**Summary Box:** *What is already known?:*

- Increasing male involvement (MI) in maternal health (MH) is considered to be a promising and effective intervention for improving maternal and newborn health outcomes.
- Male involvement is described as a multifaceted concept in the quantitative literature, although a universal definition or evidence-based set of indicators is lacking.
- In qualitative literature male involvement is often described by men and women from different settings as the male partner “being there”, meaning giving physical and emotional support.

*What are the new findings?:*

- Conceptualisation of MI in MH in the literature is done by focusing on either psychosocial aspects of MI in MH or on maternal health care utilisation. The attention given to one or both aspects resulted in the use of different indicators and depended on the geographical context of the study.
- Overall male involvement was most often measured by instrumental actions such as presence at health services, financial support or providing transport. Other aspects of male involvement, such as communication, emotional support and shared decision making have received little attention, especially in low- and middle-income countries.

*What do the new findings imply?:*

- More research into other aspects of male involvement (such as the subjective feeling of perceived support and decision making) can broaden the potential of male involvement programs and also reveal and minimize potential negative side-effects of male involvement interventions.
- A standardized definition and set of indicators, exploring different aspects of male involvement, could facilitate researchers to generate more robust findings, strengthening the existing evidence on male involvement programmes.

## Background

Since the 1994 Cairo Conference, where men’s involvement in contraception, family planning, maternal health and child health was emphasized, research has increasingly paid attention to men’s role, responsibilities and behaviour in sexual and reproductive health(1). Evidence about the positive impact of male involvement in maternal health on maternal and child health outcomes has been widely published in the last decade (2–6) and recently the World Health Organization (WHO) included active involvement of men during pregnancy, child birth and the postpartum period as an effective strategy to improve maternal as well as newborn health outcomes in their 2015 recommendations on maternal and newborn health promotion interventions (7).

However, there is no common operational definition of male involvement, nor is there a set of indicators, despite the body of evidence about the positive impact. Researchers seem to agree that MI is a multifaceted term but the concept itself has taken different forms according to the context and researcher’s interest. Looking through the lens of Prevention of Mother to Child Transmission (PMTCT) programmes for example, researchers often focus primarily on male presence at antenatal care (ANC) and HIV testing(3,8–10) as the core indicators for male involvement, without paying attention to other aspects of involvement in maternal health. This single measurement assumes that male partner presence is always a positive action and that men who do no not attend services are inherently “not involved” (11). However, it is well known that in many health systems men face multiple barriers to being present during ANC such as privacy issues, overcrowded ANC consultations, stigmatization and strong prevailing gender norms(12–14). Consequently, the fact that he is or not present might not correspond to his intentions of being involved or actual (supportive or unsupportive) behaviour outside the health facility. Limited research has also highlighted the negative side of male presence at ANC(12,15,16). In some cases it might be an act of dominance and control, thereby limiting women’s ability to actively participate in the conversation during the antenatal care consultation(12). All these arguments should be taken into account when measuring male involvement based on a single indicator.

Looking at the qualitative literature there seems to exist some consensus regarding the meaning of male involvement globally, with slightly different accents according to the context. A study in rural South Africa showed that male involvement was understood as giving instrumental support to female partners through financial help, helping with physical tasks, and providing emotional support(17). In Mozambique male involvement was seen as “taking care of the family” in various ways such as providing financial support, making the decisions and showing love towards the partner(12). In two Arabic countries male involvement was described as being accessible, present, and available in addition to being supportive and encouraging(18). Studies from the United States found that male involvement meant “being there”, both emotionally and physically, by doing household chores or listening attentively to the woman’s concerns(19,20). African American parents in the US summarised male involvement as being present, accessible, available, understanding, willing to learn about the pregnancy process and eager to provide emotional, physical and financial support(21). Despite the common construct of “being there”, often meaning supporting financially, emotionally, and being physically present, this has not yet been translated into a set of robust quantitative indicators for measuring male involvement. Nevertheless more recently studies have started to construct composites or a collection of indicators for measuring male involvement, instead of focusing on a single item(22,23). Furthermore, factors such as financial support, birth preparedness, decision making and participation in household chores have been included as male involvement indicators. Some studies also included reports of the male partner himself, often resulting in contradicting findings between men and women(24).

Despite the growing number of studies in the field of male involvement in maternal health, no consensus exists regarding the number and content of indicators for assessing male involvement in maternal health, although several authors have argued that evidence based indicators are necessary for improving the quality of the available evidence(3,16,25). With this review we want to explore to what extent the research community has assessed different dimensions of male involvement in maternal health and which patterns we can identify in the selection of indicators globally. Only by looking at male involvement through a broader lens can potential implications of male involvement interventions on different outcomes such as gender equality, psychosocial health and couples birth preparedness be explored and improved.

The primary aim of this systematic review is to examine the conceptualization of male involvement in maternal health in the quantitative literature of the last twenty years. As secondary objective we want to critically review and discuss commonly used indicators.

## Methodology

### Protocol and registration

This systematic review was conducted in accordance with the PRISMA (Preferred Reporting Items for Systematic Reviews and Meta-Analyses statement of 2015) guidelines. The protocol was published online on the 10^th^ of July 2020 on PROSPERO (International prospective register of systematic reviews) under registration number CRD42020169078.

### Eligibility criteria

The systematic review included all types of quantitative studies involving indicators or variables representing male involvement in maternal health published in the last 20 years. Male involvement was defined as the participation, engagement or support of men in all activities related to maternal health. Maternal health was defined according to WHO as the period from conception until 6 weeks after childbirth, thus covering pregnancy, childbirth and the postpartum period.

A search strategy was developed by the principal investigator (PI) with inputs from ODG. This search was refined with the help of the librarian of Ghent University. The systematic review involved a literature search of the PubMed, Embase, Scopus, Web of Science and CINAHL databases for peer-reviewed journal articles. Grey literature was identified through the WHO Reproductive Health Library and using Google with relevant keywords. The search strategy for Pubmed can be found in additional file 1. A final search was conducted on the first of May 2020. The outputs of the search were exported to Mendeley desktop V.1.16.1, and duplicates were removed. Subsequently, the titles and abstracts of the studies were imported into Rayyan. Two rounds of screening were applied, first, by title and abstract, followed by full text. Two reviewers independently screened and appraised all eligible articles using pre-set criteria, and in case of disagreement consensus was reached through discussion. Exclusion criteria were: using only qualitative methodology, systematic review studies, conference abstracts, data collection limited to postpartum period and not during pregnancy or childbirth, articles without any measurement of the role of the partner, and studies limited to testing of the male partner for HIV or STIs without mentioning male support, involvement, engagement or participation.

In the second stage of screening full texts were obtained for the screened abstracts. If the article was unavailable through an online search, the article reach system of Ghent University was used to obtain the articles or the authors were contacted to request the full text publication. The same criteria were applied for inclusion and exclusion as in the first stage of screening together with a quality appraisal.

### Quality assessment

Papers selected for retrieval in the second stage were assessed by two independent reviewers for methodological validity prior to inclusion in the review, using standardized critical appraisal instruments from the Joanna Briggs Institute (including the checklist for analytical cross sectional studies, cohort studies, prevalence studies, quasi-experimental studies, randomized controlled trials, case control studies and systematic reviews). Studies with a score below 50% were excluded. A relatively low threshold was used for inclusion because we wanted to examine the concept of male involvement used by the wider scientific community on a global scale, rather than limiting our results to a few high quality studies.

### Data extraction and analysis

A pre tested data extraction framework in Microsoft Excel was used to extract and chart data from the reviewed articles. The standard data extraction table included authors, publication year, topic of study, the exact term used for describing male involvement (involvement/engagement/support/participation), year of study, study design, geographical location of study, definition of male involvement (if given), indicators used for measuring male involvement, data sources, and quality assessment. Only indicators used in more than five of the included studies were retrieved for the results section and studies referring to a scale of more than 10 items (always psychosocial scales) were categorized as “Psychosocial scale measurement (>10 items)”. Data extraction of every article was done by a team of three researchers. The PI (AG) screened all articles and GP and TVS independently each screened 50% of the articles. Disagreement (<10%) was resolved by discussion or consultation with one of the supervisors (ODG) if needed. Topic allocation was done by manually giving the article one of the following topics as the “core topic”: prevention of mother to child transmission (PMTCT), psychosocial health (PSH), abortion, and maternal and newborn health (MNH). Articles were categorized as PMTCT if they focused on care for women living with or at risk of HIV (to maintain their health and prevent transmission to their babies), including studies focusing on male HIV testing and prevention. PSH categorized studies focused on social and emotional aspects of male partners’ role in maternal health, mainly consisting of articles regarding perinatal depression and stress. Studies categorized as abortion focused on women considering or having experienced an induced or spontaneous abortion. The topic MNH included all studies focusing on male involvement in maternal and newborn health, excluding the previous categories (PMTCT, PSH or abortion). The topics were inductively created and discussed until agreement was reached among the three reviewers responsible for data extraction. The data of the data extraction sheet was cleaned and subsequently analysed using descriptive analysis. Fisher’s exact test was used for assessing differences in the main topic according to terminology and according to continent. In addition, confidence intervals were calculated for visualizing differences in proportions in the in use of indicators according to terminology.

For visualization and confirmatory analysis of the data from the included articles, text mining by R with the tidytext package was used for natural language processing (26). First a test set of 20% of the data was used for writing the text mining scripts, which were refined once the full dataset was entered. The statistic “term frequency-inverse document frequency” (tf-idf) in combination with n-grams was calculated for assessing the importance and structure of certain word combinations within the collection of articles (referred to as “corpus”). A word’s *tf-idf* represents the frequency of a term adjusted for how rarely it is used. The statistic *tf-idf* is intended to assess how important a word is to a subset of documents in a collection of documents or corpus(26).

Lastly, Latent Dirichlet Allocation (LDA) was used for Topic modelling, which is a mathematical method for finding clusters of words that characterize a set of documents(26). Latent Dirichlet Allocation is a commonly used algorithm for topic modelling in text mining and was used in our study to identify meaningful topics within the complete set of included articles, our so called “corpus”(26). An essential step of the LDA algorithm is assigning each word in each article to a topic. The more words in a document are assigned to that topic, generally, the more weight (or gamma probabilities) will go on that document-topic classification.

## RESULTS

Electronic database searches identified 5277 titles and abstracts, with a further 7 identified through the grey literature search. After removal of duplicates, 3975 articles were screened by title and abstract, resulting in 569 potential articles to be included. After reviewing the full text of these articles, 282 unique studies were included in the systematic review. A flowchart regarding the inclusion of articles can be found as an additional file (additional file 2).

### Characteristics of included studies

Of all included studies, most studies were conducted in Africa (43%), followed by North America (23%), Asia (15%) and Europe (12%) (see table 1). The majority of studies collected data from women only (58%), while 20% collected data from both men and women and around 16% collected data from men only. Registry data was used in six percent of studies and mostly referred to hospital files indicating the presence of men during ANC. Terms used to assess the role of the male partner were: involvement, support, engagement, participation, attendance and presence. Most studies used a cross sectional design (58%), followed by a longitudinal design (23%). Only around six percent of the studies used a randomized controlled trial design. One in ten studies did not give a clear definition of the indicator used for assessing male involvement (or one of the similar terms listed earlier). Most studies (63%) used a combination of indicators for measuring male involvement. Two fifths (40%) of studies used ANC attendance as one of their indicators for assessing male involvement and one in six studies used financial support or transport (17%) and presence during delivery (17%). Around 14% of studies used the indicator HIV testing. All indicators that were used to assess male involvement can be found in Table 1. Core topics of the studies were psychosocial health (32%), maternal and newborn health (MNH) (48%), PMTCT (18%) and abortion (3%). All categories within Table 1 were mutually exclusive except for the used indicators;

**Table 1.**
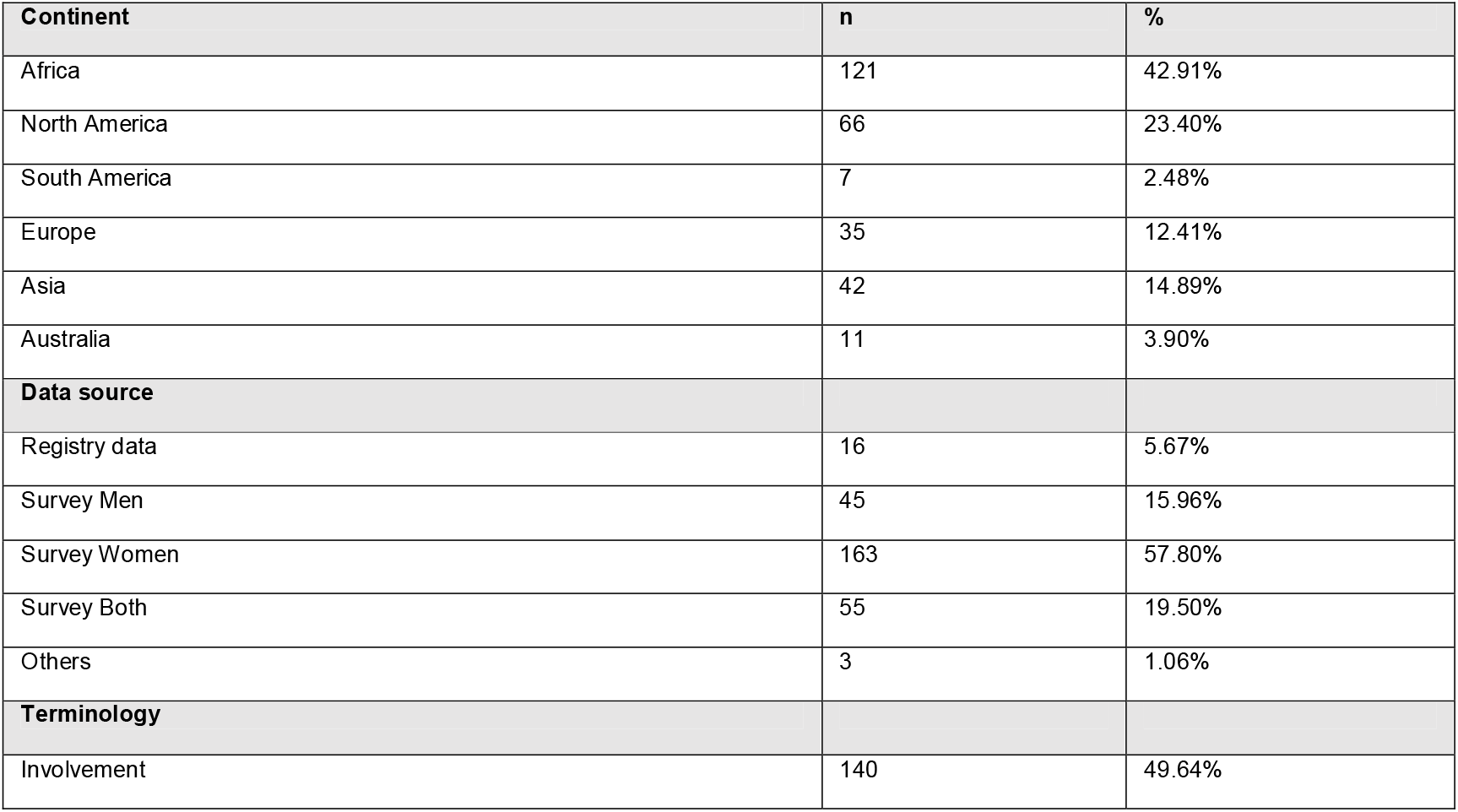
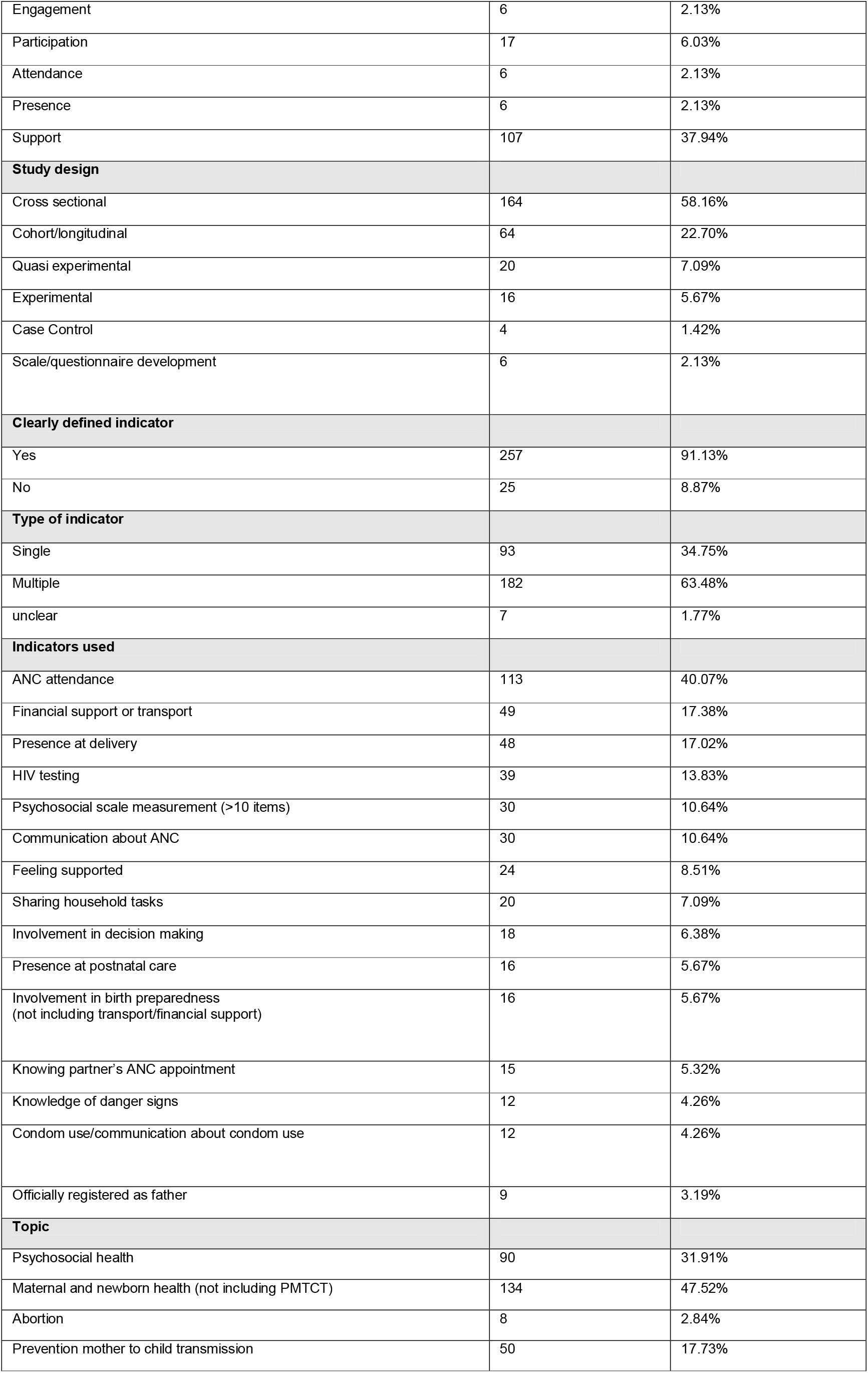
Characteristics of the included studies.

| Continent | n | % |
| --- | --- | --- |
| Africa | 121 | 42.91% |
| North America | 66 | 23.40% |
| South America | 7 | 2.48% |
| Europe | 35 | 12.41% |
| Asia | 42 | 14.89% |
| Australia | 11 | 3.90% |
| Data source |  |  |
| Registry data | 16 | 5.67% |
| Survey Men | 45 | 15.96% |
| Survey Women | 163 | 57.80% |
| Survey Both | 55 | 19.50% |
| Others | 3 | 1.06% |
| Terminology |  |  |
| Involvement | 140 | 49.64% |
| Engagement | 6 | 2.13% |
| Participation | 17 | 6.03% |
| Attendance | 6 | 2.13% |
| Presence | 6 | 2.13% |
| Support | 107 | 37.94% |
| **Study design** |  |  |
| Cross sectional | 164 | 58.16% |
| Cohort/longitudinal | 64 | 22.70% |
| Quasi experimental | 20 | 7.09% |
| Experimental | 16 | 5.67% |
| Case Control | 4 | 1.42% |
| Scale/questionnaire development | 6 | 2.13% |
| **Clearly defined indicator** |  |  |
| Yes | 257 | 91.13% |
| No | 25 | 8.87% |
| **Type of indicator** |  |  |
| Single | 93 | 34.75% |
| Multiple | 182 | 63.48% |
| unclear | 7 | 1.77% |
| **Indicators used** |  |  |
| ANC attendance | 113 | 40.07% |
| Financial support or transport | 49 | 17.38% |
| Presence at delivery | 48 | 17.02% |
| HIV testing | 39 | 13.83% |
| Psychosocial scale measurement (>10 items) | 30 | 10.64% |
| Communication about ANC | 30 | 10.64% |
| Feeling supported | 24 | 8.51% |
| Sharing household tasks | 20 | 7.09% |
| Involvement in decision making | 18 | 6.38% |
| Presence at postnatal care | 16 | 5.67% |
| Involvement in birth preparedness<br>(not including transport/financial support) | 16 | 5.67% |
| Knowing partner's ANC appointment | 15 | 5.32% |
| Knowledge of danger signs | 12 | 4.26% |
| Condom use/communication about condom use | 12 | 4.26% |
| Officially registered as father | 9 | 3.19% |
| **Topic** |  |  |
| Psychosocial health | 90 | 31.91% |
| Maternal and newborn health (not including PMTCT) | 134 | 47.52% |
| Abortion | 8 | 2.84% |
| Prevention mother to child transmission | 50 | 17.73% |

### Terminology used for describing the role of the male partner

Studies using the term “male attendance”, “male participation” or “male engagement” to describe the role of the male partner did not significantly differ from studies using the term “male involvement” in the use of the most common indicators (ANC attendance, Financial support or transport, Presence at delivery, HIV testing, Psychosocial scale measurement and communication about ANC). Results of the Fisher exact test per indicator can be found in additional file 3. We consider these terms as synonyms for the remaining results section.

Studies using “male presence” to describe the role of the male partner always used presence at delivery as their only indicator and differed significantly (P=0.034) in the use of indicators from studies using the term male involvement.

Studies using the term “male support” to describe the role of the male partner showed a significant difference (P<0.001) in the use of indicators compared to the term “male involvement”. Studies referring to the role of the partner as “partner support” used more often complex psychosocial scales (>10 items) such as the Tilburg Pregnancy distress scale or Social Support Effectiveness questionnaire.

A comparison between the use of indicators for the terms using involvement/engagement/attendance or participation versus support with the respective confidence intervals can be found in Figure 1. Studies using the term “male presence” were excluded from this specific analysis because they always used presence at delivery as single indicator for MI, resulting in 276 included studies (n=276).

**Figure 1.**
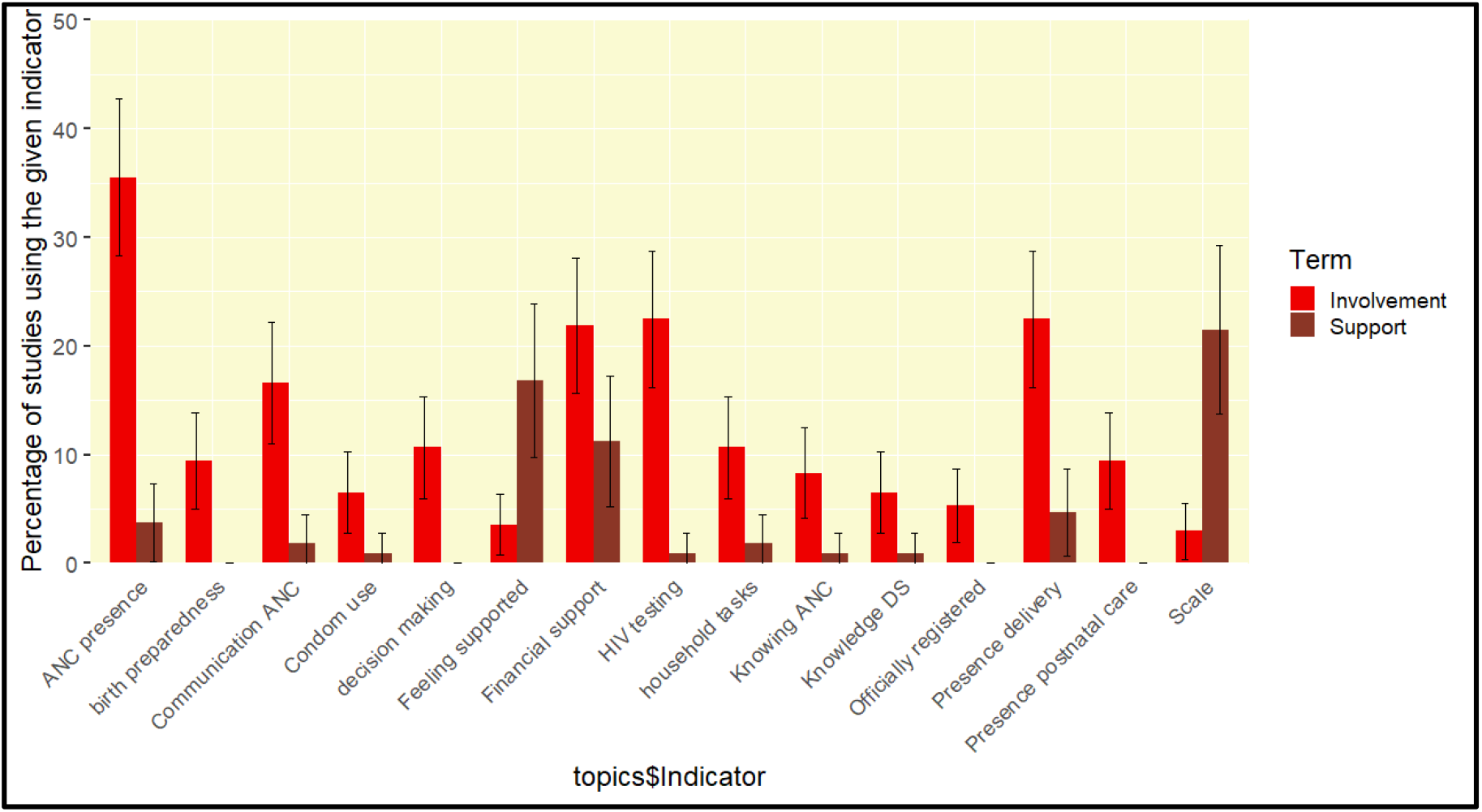
Use of indicators according to the term involvement (/engagement/attendance/participation) versus support (n=276)

### The use of different indicators for assessing the role of the male partner

Among all studies using the term male involvement/engagement/attendance or participation, a wide range of indicators was used, often assessed in different ways. On the one hand, we found studies aiming for an in-depth comprehensive assessment of male involvement through the use of extended surveys for both men and women combining different indicators (>3) for assessing male involvement (n=26). On the other hand, we found studies assessing male involvement very simply and using only service registry data retrospectively, defining male involvement as having the father’s name written on the birth certificate (n=7). In conclusion, no common set of indicators could be identified for measuring male involvement/engagement/attendance or participation.

ANC attendance as a single indicator (n=26) was common in studies in low income countries (LICs), with majority of studies deriving from Africa, Asia and South America and only two studies from North America. On the other hand, presence at delivery was used as an indicator in all continents, suggesting it is a more “universal” indicator. Financial support, which was used in all continents except in South America, was also used frequently globally as a MI indicator. HIV testing was a typical indicator in African countries and the use of psychosocial scales was more common in North American and European studies.

We were also able to identify some patterns when we examined which studies used less common indicators and why. Studies defining male involvement by having a father/partner registered on birth certificates (n=9) were most often conducted in North America (six out of nine studies), using big datasets and focusing on neonatal health outcomes. Studies focusing on knowledge of danger signs (n=12) were typically derived from low income countries (11 out of 12 studies) and focussed on maternal health outcomes (11 out of 12 studies). Studies using condom use (or communication about condom use) as an indicator of male involvement always derived from an African country and focused in the majority of cases on PMTCT (8 out of 12 studies).

### The relationship between terminology, continent of the study and topic

A scatter plot showing the relationship between the continent of the study, the topic of the study and the terminology can be found Figure 2. A significant difference was found in the term used in the study according to continent (p<0.001) and topic (p<0.001). Studies using the term “support” were more often conducted in Europe, North America, Australia and Asia, while the term “involvement” was most often used in Africa and South America. All continents had studies using both terms.

**Figure 2.**
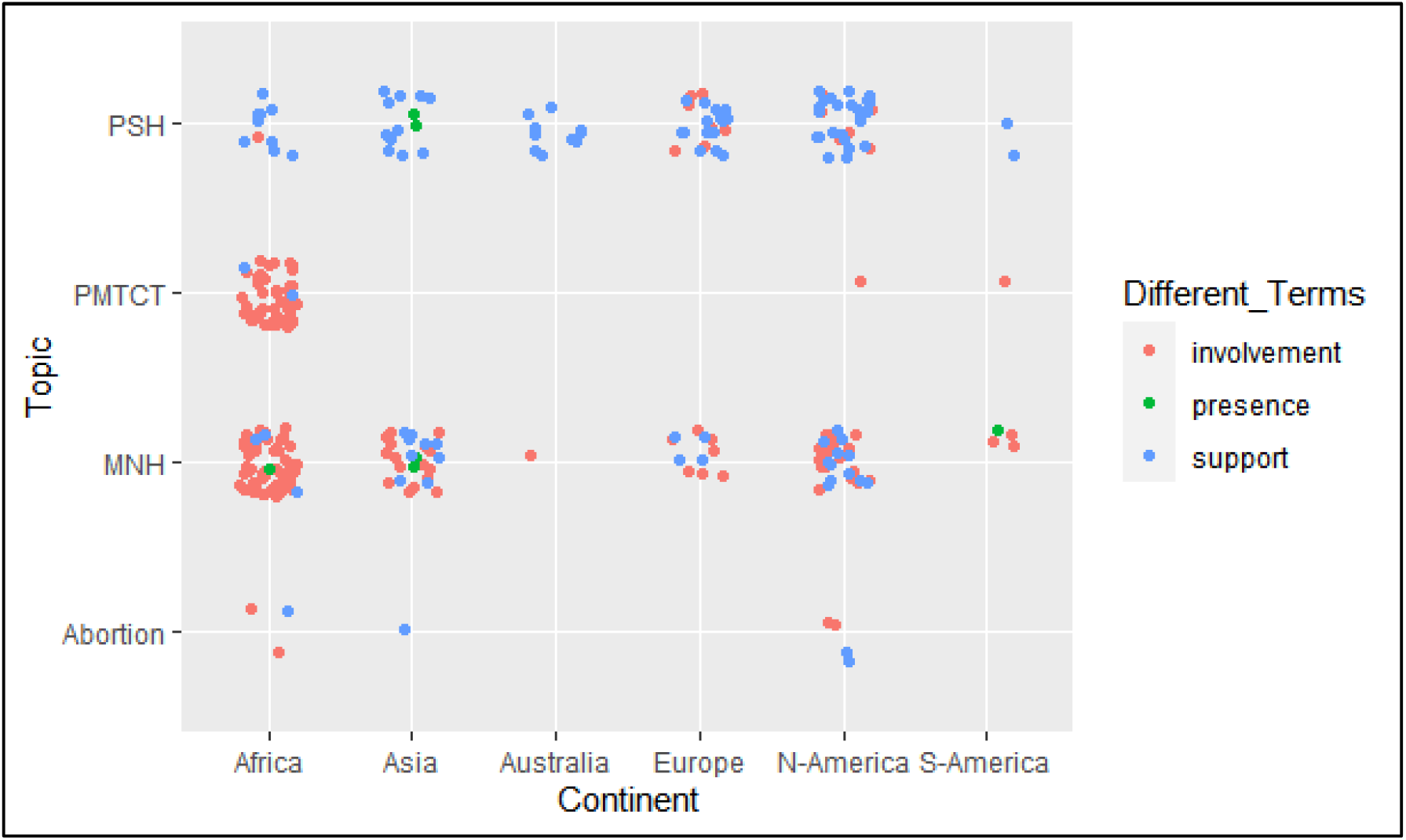
Jittered scatterplot showing the relationship between the manually classified topic, continent and terminology of the different studies included in the review (n=282); PSH=Psychosocial Health; PMTCT=Prevention Mother To Child Transmission; MNH= Maternal and Newborn Health.

Looking at the main topic of the study (manually given during data extraction) we also found a significant difference according to continent (p<0.001). The topic “prevention of mother to child transmission” was most prevalent in Africa while the topic “psychosocial health” was common for studies from Europe and Australia (see Figure 2).

In line with the indicators used for measuring male support (which mostly used psychosocial scales), studies using the term “support” also more often had as their main topic “psychosocial health” (p<0.001) (see Figure 2).

### Text mining: highly used words according to continent

Pairs of two consecutive words, referred to as “bigrams”, were examined by text mining using the tidypackage. The tf-idf statistic was calculated for the bigrams in the corpus. Subsequently the top 15 words were ranked per continent. The continents Australia and South America were deleted during the process because of their low number of articles, 11 and 7 respectively. In Figure 3 the top 15 words per continent are represented in a word cloud, with font size reflecting the tf-idf value, showing the different content of the articles according to continent. The word cloud shows that in Asia male involvement studies were characterised by a focus on nutrition (reflected by the words “maternal nutrition”, “healthy moms”, “disordered eating” and “added sugar”). In Africa, institutional delivery (reflected by the words “skilled delivery”, “supervised delivery” and “delivery site”) seem important, as well as PMTCT and birth preparedness. In North America the words “relationship stress”, “IPV” (intimate partner violence) and “PPD” (post-partum depression) show that the literature mainly focuses on psychosocial health and the couple relationship. “Abortion” and “adolescents” were more prominent words in the literature in North America compared to other continents.

**Figure 3.**
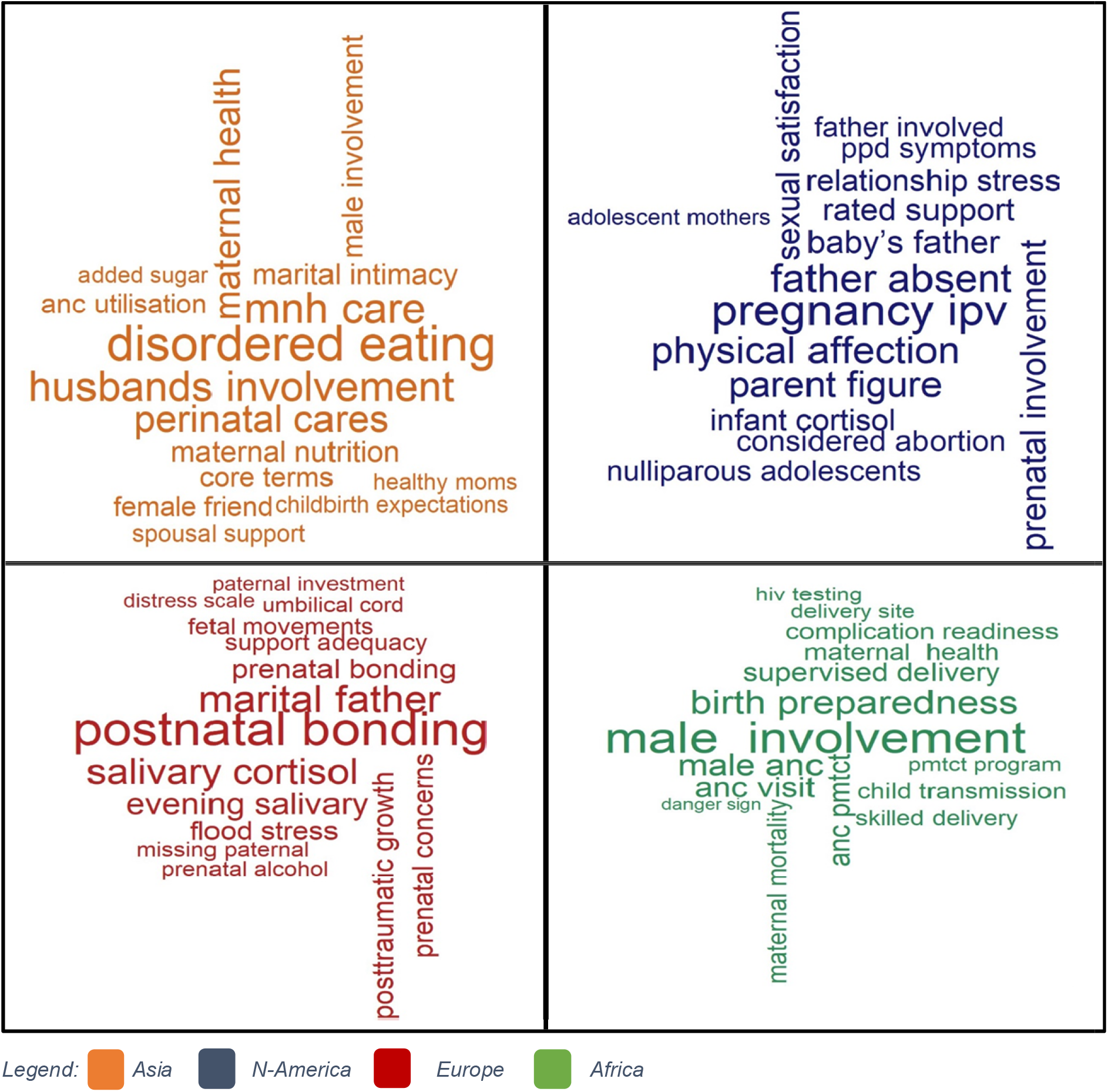
Word cloud visualising the top 15 bigrams per continent, based on their tf-idf value.

### Latent topic allocation by text mining

Lastly, we conducted a latent Dirichlet topic allocation by setting k = 2, to create a two-topic LDA model. A larger number of topics (i.e. a larger k) resulted in an unclear pattern of words, which were difficult to interpret. Setting k=2 we could find two meaningful topics in the corpus. First the algorithm identified two topics in the corpus and subsequently we calculated the probability that a word corresponded to topic 1 and to topic 2 (by gamma probabilities) and similarly the probability that a complete document (or article) belonged to topic 1 and topic 2.

Figure 4 is a scatter plot of the words that are most common within each topic (with the respective gamma probabilities on the x-axis and y-axis) and diagonal lines at 30 degrees and 60 degrees for separating the words that are common among both topics. The scatter plot shows the two different meanings of the two topics that were extracted from the articles. The most common words in topic 1 include “HIV”, “PMTCT”, and “testing”, suggesting that this topic represents the topic of HIV prevention, but also more broadly antenatal care attendance (by words such as attendance, visit, facility, clinic). The most common and meaningful words in topic 2 include “support”, “depression”, “stress”, “social” and “psychological”, which suggests it may represent studies around psychosocial health. Furthermore topic 2 includes the word “father”, while topic 1 only includes the word “male”. This might indicate that studies in the field of psychosocial health have a longer follow up and more often include the period after birth, when the male partner has become a father. The words between the diagonal lines are common among both topics and include words such as “women”, “pregnant”, “maternal” and “child”. This is an advantage of topic modelling as opposed to “hard clustering” methods: topics used in natural language could have some overlap in terms of words.

**Figure 4.**
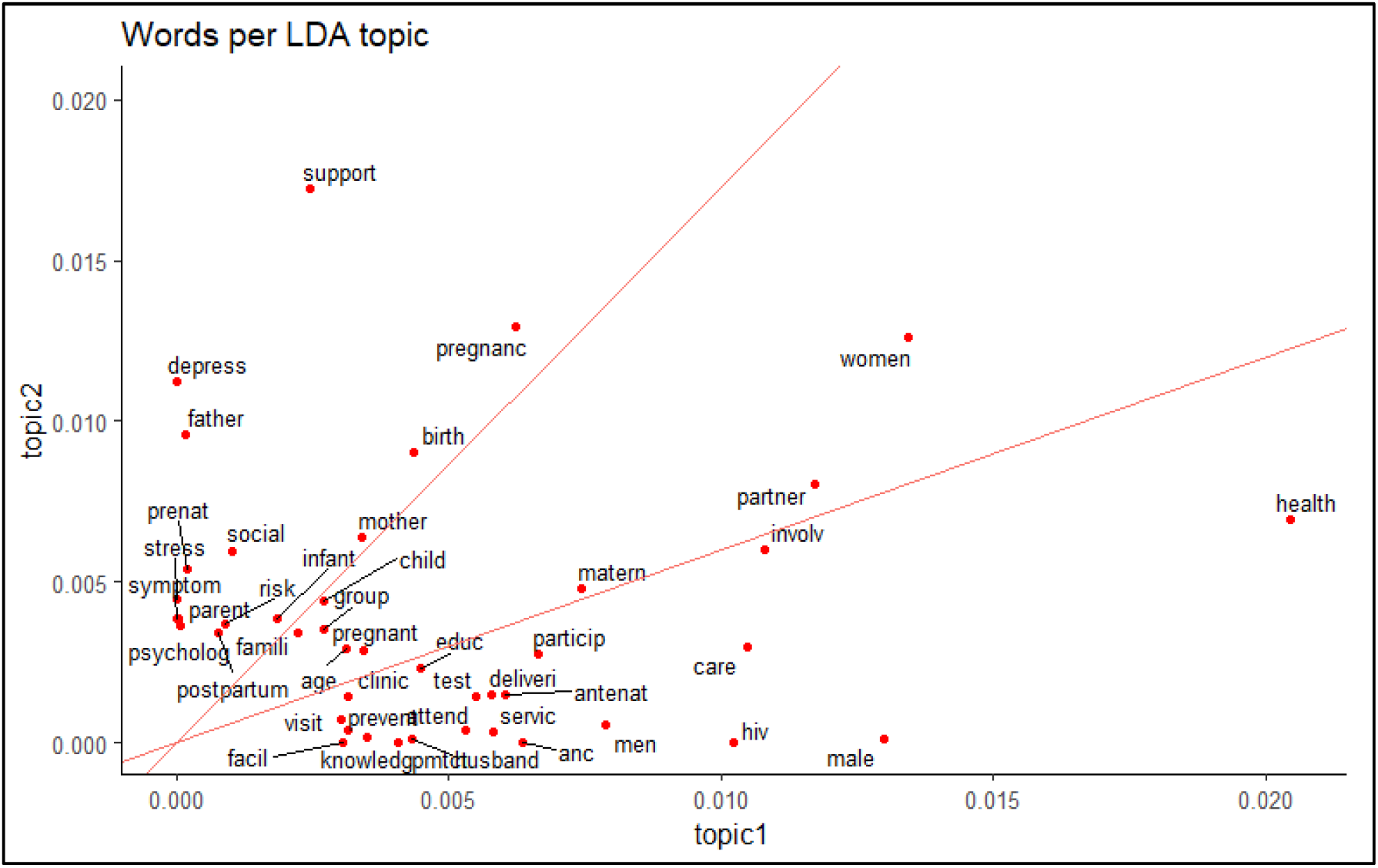
Most common words within each topic and their gamma probabilities with a diagonal line at slope 1.73 (30°) and 0.60 (60°)

### The relationship between the computer driven topic allocation, manually classified topics and continent of the studies

After identifying these topic probabilities per word, we explored how well our unsupervised learning did at distinguishing the different topics in the documents. We would expect that studies that were manually given the topic PMTCT would be found to be mostly (or entirely) part of LDA topic 1 (HIV prevention and antenatal care attendance) and that studies given the topic “psychosocial health” would correspond to LDA topic 2 (psychosocial health). In addition, we were interested in the extent to which the topic “abortion” and “MNH” would correspond to one or both of the LDA topics.

We visualized the relationship between the LDA Topics, manual topics, and continents by a scatter plot (see Figure 5) and calculated the associations between LDA topics and manual topics with a Cross Tabulation. The latter demonstrated that the topic PMTCT highly corresponds to LDA topic 1 (HIV prevention and antenatal care attendance), with 92% (46/50) of the PMTCT studies being classified as LDA topic 1. The topic PSH strongly corresponded to LDA topic 2 (psychosocial health) with 92% (83/90) of the PSH studies classified as topic 2. For the topic “abortion” we saw a higher correspondence with topic 2 (75%; 6/8) (psychosocial health) and lower correspondence with topic 1 (25%; 2/8). The manual topic MNH seemed to correspond to LDA topic 1 (61%; 82/134) and 2 (39%;52/134). In the scatter plot (see Figure 5) we noted a remarkable difference between LDA topic and manual topic correspondence according to continent, whereby MNH seem to correspond to topic 1 in Africa (HIV prevention and antenatal care attendance) but to topic 2 in North America (psychosocial health). This indicates that while several studies in North America received the label “MNH” during our manual data extraction, the natural language of studies in North America clearly differs from those conducted in Africa, resulting in another LDA topic (related to psychosocial health).

**Figure 5.**
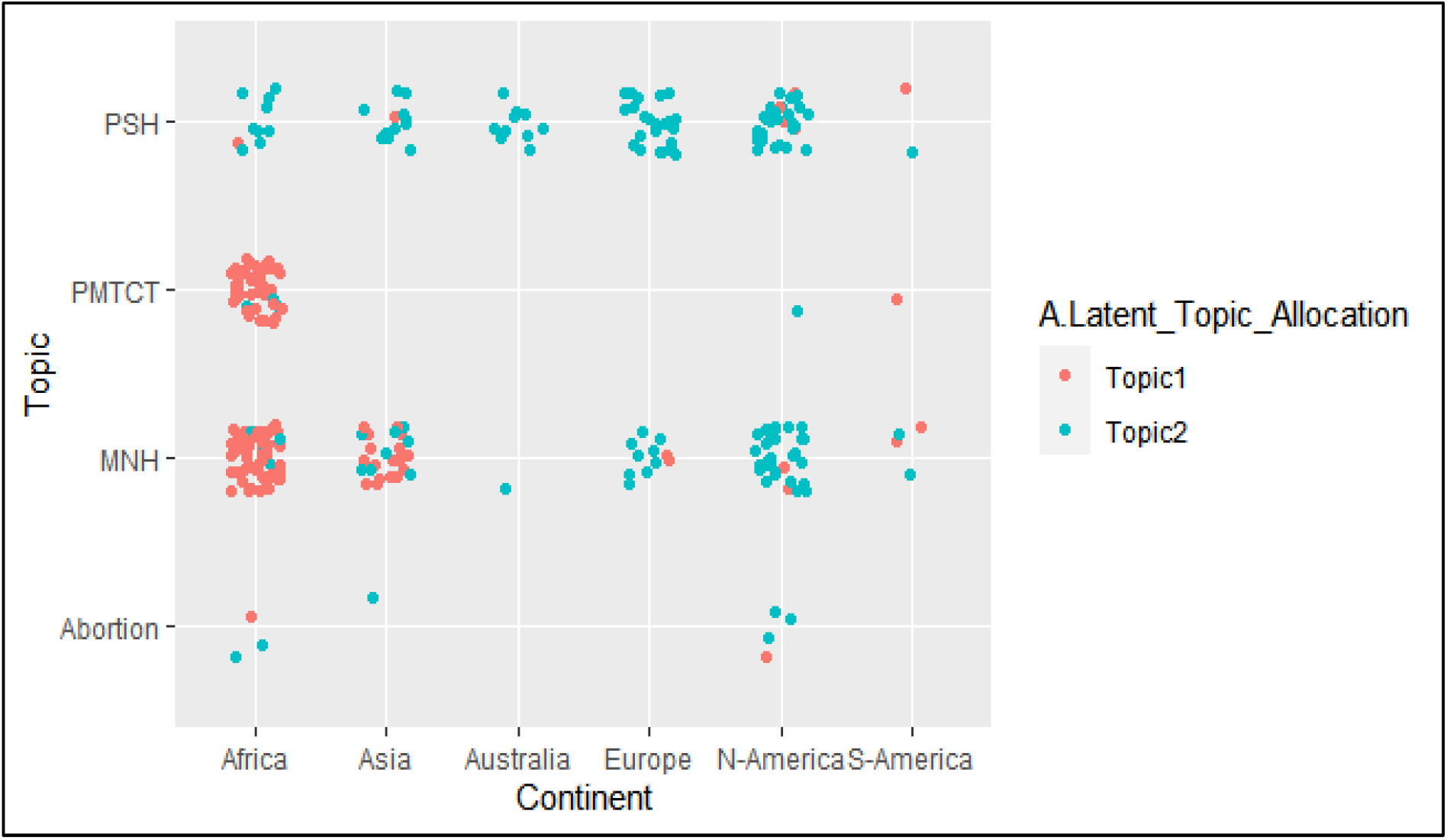
The relationship between the manual topics, LDA topics and continent of the included studies (n=282); PSH=Psychosocial Health; PMTCT=Prevention Mother To Child Transmission; MNH= Maternal and Newborn Health.

## Discussion

The broad range of studies in the scientific literature examining and assessing male involvement have formed the evidence base for promoting male involvement as a promising strategy for improving maternal and newborn health outcomes(2,27,28). With this systematic review we aimed to examine the conceptualization of male involvement in maternal health in the quantitative literature of the last twenty years and critically review and discuss commonly used indicators.

Both manual and computer driven topic allocation showed us that studies in the field of male involvement in maternal health are mostly conducted to examine psychosocial health on the one hand and maternal health care utilisation (especially ANC attendance, PMTCT services and institutional childbirth) on the other hand. Looking at geographical context, several differences were noted by focusing on the most unique and common terms using text mining. In studies deriving from Asia words related to nutrition were more important, while in studies from North America words referring to intimate partner violence were more typical. In studies conducted in Europe, stress and depression were important terms, while in Africa antenatal care attendance and HIV prevention were important. Some of these differences can be explained by the different prevalence of certain problems (such as malnutrition in India(29) and HIV in Africa(30)), while other differences are less logical and probably influenced by funding bias and geographical sociocultural factors. Future research could explore new pathways in male involvement studies (such as the relationship between psychosocial health and male involvement in an African context), building on the evidence from other countries and continents.

A very low number of studies derived from South America compared to Africa, indicating male involvement might not be considered a high priority issue in maternal health care in that region. However, the limited literature emphasizes that the current level of male involvement is extremely low in South America(31,32), with strong gender norms being the most persistent barrier(33,34). The reason why until now very few programmes have focused on male involvement in South America might be related to the lower prevalence of HIV on the continent compared to Africa(30). Historically many male involvement programs in maternal health in LMIC were implemented in order to improve the uptake of PMTCT, making it less useful (and less funded) in countries with low HIV prevalence. This also explains why the literature on male involvement in South America is more often focused on attaining gender equality instead of getting men to health facilities for HIV testing (35,36).

The proportion of studies about psychosocial health and depression was lower in Africa compared to other continents. However, the literature indicates that perinatal depression is common in the African region(37–40). Globally, perinatal depression is estimated to affect around 11% of women, and recent studies have shown that perinatal mental disorders (depression and anxiety) are at least as prevalent in Africa as in other regions (41–44). Furthermore, research has demonstrated that HIV positive women have increased risk of perinatal depression. A systematic review found a prevalence of 23.4% for antenatal depression and 22.5% for postnatal depression in HIV infected women(45,46). The low number of studies on psychosocial health in Africa in our review showed that the relationship between perinatal depression/psychosocial health and the role of the male partner is poorly studied in Africa. While some have argued that many low income countries have more pressing issues within maternal health than addressing perinatal depression (such as severe maternal morbidity and mortality)(2), other studies have shown there is reasonable evidence for the benefits and effectiveness of psychological interventions in LMIC(44,47). The low availability of mental health services in LMICs is one of the main challenges for addressing mental health problems, but some recent studies have shown that training and organising lay mental health care workers to address mental health care problems are a feasible and effective approach to combatting mental health disorders(48). Furthermore male involvement and/or partner support has been shown to be a protective factor against perinatal depression globally(49–52). More research into the field of male involvement and maternal mental health could provide multiple health gains for the male partner, mother and child in LMICs.

Very few studies (n=8) focused on the role of the partner during abortion care. Unintended pregnancies and access to safe abortion services are major public health concerns, in which the important role of the partner in decision making and access to services cannot be ignored (53,54). A systematic review from 2016 showed that women contemplating abortion frequently involve their male partner in the decision and rely on him to help with logistics, finances and emotional support before and after the abortion; furthermore, male involvement was positively associated with women’s well-being(54). Given that 121 million unintended pregnancies occur each year with 61% ending in an abortion, the role of the partner during abortion care seems to be under-studied within the scientific literature (53).

In general, the benefits of father involvement for the father himself are hardly studied. Most studies in our review did not collect data directly from men and even fewer studies assessed the potential benefits of male involvement for the father himself or mutual perceived support. Father involvement is almost always used as an instrumental approach to improve maternal health, although the added value for the father himself (for example the perceived health benefits by improving his own access to health care services) were already highlighted during the Cairo conference in 1995 (55). Emphasising the positive effect for the father himself could be explored as an intervention strategy(56), whereby the health benefits might go beyond his participation in maternal health care services.

Despite the consensus that male involvement is a multifaceted concept, majority of studies seem to focus on only one particular aspect of the concept, resulting in a simplified measurement of male involvement in maternal health. The latter was illustrated by the high number of studies relying on a single indicator. Furthermore, there was no common set of indicators among studies using a combination of different indicators, almost every study had its own unique composite. Obviously, the measurement of male involvement depends on the context, but a critical reflection of the measurement is needed for a correct interpretation of the results. This is especially important as the lack of agreement in indicators leads to the risk that researchers only report the most significant variable. In some studies, included in the review, we found that male involvement was described and defined as a multidimensional concept in the introduction and methods but that in the results section only one indicator was used as “the male involvement indicator”. As a consequence results might be biased by selecting and reporting only the most significant indicator.

Almost half of the studies focused on presence at antenatal care or HIV testing and consequently the benefits for mothers and their newborns will mainly be oriented towards the prevention of HIV transmission. This coincides with the implementation of instrumental male involvement policies in several countries, aiming at improving male attendance at ANC by refusing to attend women without a partner present or giving priority to couples in the waiting line(12,57–59). The negative side effects of introducing such policies for improving male attendance at ANC have started to emerge (such as increased gender inequality, stigmatisation of single women and lower ANC attendance of women (12,60)), nevertheless they have not led to the elimination of such programs. This might be related to the strong influence of HIV programmes and donors, where programme success is defined by the proportion of men being tested during pregnancy(61). In many communities men attending antenatal health care services are perceived as being HIV positive (62), because historically HIV counselling and testing was the main reason for inviting men in several African countries(61). Future male involvement programs should try to shift away from the focus on HIV testing and break the circle of stigmatisation that has been associated with these programmes.

Certain aspects of male involvement such as communication, decision making and “feeling supported” were rarely included as male involvement indicators (10.64%, 8.51% and 6.38% respectively) in the studies included in our review, while both quantitative and qualitative research have shown that these aspects of men’s involvement play an important role in maternal health care access, utilisation and outcomes (63–68). The narrow focus on specific actions of men (such as financial support and ANC attendance) without taking into other aspects (such as couple dynamics) clearly entails a risk of missing essential information and underreporting negative consequences. More data collection regarding empowerment of women, gender equality and perceived support (from both men and women) is needed for assessing potential side effects of male involvement programs (13,69). A limited number of studies has shown that interventions could unintentionally lead to increased domination of decision-making about pregnancy, nutrition and infant care by men, putting pressure on women to adopt certain beliefs and practices(4,70) (71). Furthermore disclosure of IPV during antenatal care might be hampered by male involvement strategies due to a lack of privacy(72,73). Only by aiming for a comprehensive assessment of male involvement programs (collecting both quantitative and qualitative data) can these issues be identified and addressed in future interventions. Lastly, the collection of sociodemographic data could be essential for correct interpretation of the data, especially when male involvement is assessed by a single indicator. Some types of involvement might be hampered by poverty (such as providing financial support or transport) or other factors (for example, men working as seasonal workers away from home will not be present at ANC). Only by taking into account all these factors can effective programs be set up, leading to improved maternal and newborn health outcomes (including psychosocial health) and improved health and wellbeing outcomes for the father himself, as well as increased gender equality. In conclusion we believe the evidence base on male involvement indicators needs to be improved in the future in terms of regional representation, study robustness, and a broader holistic scope.

### Limitations

This review has certain limitations. We only included quantitative studies and used qualitative literature only for interpretation of the results. A similar in-depth systematic review regarding the qualitative meaning of male involvement, comparing findings from different regions, would complement our findings. Furthermore, a broader review also including the role of the partner in planning a pregnancy (before conception) and/or family planning decisions could strengthen the existing evidence regarding male involvement in reproductive health.

Another bias in this study might be related to the PI’s background. A researcher’s background and position often affects what they choose to investigate, the angle of investigation, the methods judged most adequate for this purpose, the findings considered most appropriate, and the framing and communication of conclusions(74). AG has mainly conducted research in Mozambique, which might lead to a higher interest in the findings most relevant to this context (for example the relationship between HIV programs and male involvement). However, by involving co-authors in all stages of the research process we tried to minimise this bias. Finally, we only included English literature and restricted our search to a selected number of databases, which means certain studies will have been missed.

## Conclusion

The concept of male involvement in maternal health is considered to be a multifaceted concept but a general consensus regarding the actual meaning is missing. We found two main streams of conceptualisation within the literature: a focus on psychosocial support on the on hand and focus on instrumental support for maternal health care utilisation (such as PMTCT services, antenatal care attendance and institutional childbirth) on the other hand. While both aspects are considered as core elements of male partner’s (potential) role in maternal health, majority of studies seem to focus on only one of both aspects. In line with these findings the concept of MI in MH it is often measured by a simplified and narrow set of indicators and several essential elements such as communication between the couple regarding maternal health care issues, shared decision making, participation in household tasks and the subjective feeling of being supported by the male partner have received little attention. Further research, involving experts and pilot testing, is recommended to develop consensus regarding a robust set of valid and feasible indicators for assessing male involvement in maternal health globally in a more comprehensive way.

## Data Availability

Data analysed during the current study will be made available from the corresponding author upon reasonable request.

## Abbreviations

ANC: Antenatal Care
LDA: Latent Dirichent Allocation
Tf-Idf: Term Frequency-Inverse Document Frequency
LICs: Low Income Countries
LMICs: Low and Middle Income Countries
MNH: Maternal and Newborn health
PI: Principle Investigator
PMTCT: Prevention of Mother To Child Transmission
PSH: Psychosocial Health
WHO: World Health Organisation

## Acknowledgments

We would like to thank the librarian of Ghent University, Miss Nele Pauwels, for organising training on conducting a systematic review and further personal assistance in developing the search strategy. Furthermore, we would like to thank the colleagues of ICRH-Belgium for their continuous support and encouragement during the study.

## Contributorship

AG conceptualised the study and lead the process of reviewing the literature. GP and TVS screened articles and contributed to data extraction. AG conducted all analysis under supervision of ODG. All authors contributed to the development of the manuscript and read and approved the final version. The corresponding author attests that all listed authors meet authorship criteria and that no others meeting the criteria have been omitted.

## Ethical approval

Not required.

## Patient and Public Involvement

No patient or public involvement took place in the design or conduct of this study.

## Dissemination to participants and related communities

The authors intend to disseminate this research through social media, press releases, and media departments and websites of authors’ institutions.

## Transparency

The guarantor affirms that this manuscript is an honest, accurate, and transparent account of the study being reported; that no important aspects of the study have been omitted; and that any discrepancies from the study as planned (and, if relevant, registered) have been explained.

## License for publication

The Corresponding Author of this article contained within the original manuscript, which includes any diagrams & photographs within, has the right to grant on behalf of all authors and does grant on behalf of all authors, a licence to the BMJ Publishing Group Ltd and its licencees, to permit this Contribution (if accepted) to be published in the BMJ and any other BMJ Group products and to exploit all subsidiary rights, as set out in our licence set out at:http://www.bmj.com/about-bmj/resources-authors/forms-policies-and-checklists/copyright-open-access-and-permission-reuse.

