## Additional File 1 for "A systematic review of the concept “male involvement in maternal health” by natural language processing and descriptive analysis"

***Additional File 1: Search strategy Pubmed***

***##Concept maternal health in Mesh Terms***

((("Maternal Health"[MESH] OR "Maternal Health Services"[MESH] OR "Pregnancy"[MESH] OR "Pregnancy Complications"[MESH] OR "Delivery, Obstetric"[MESH] OR "Hospitals, Maternity"[MESH] OR "Infectious Disease Transmission, Vertical"[MESH] OR "Breast feeding"[MESH] OR "Postpartum period"[MESH] OR "perinatal care"[MESH] OR "midwifery"[MESH] OR "mothers"[MESH] OR "infant, newborn"[MESH] OR "prenatal care"[MESH] OR "postnatal care"[MESH] OR "Reproductive Health Services"[MESH] OR "Maternal Welfare"[MESH] OR "Abortion, Induced"[Mesh])

***##Concept maternal health in TIAB***

OR (maternal[tiab] OR abortion[tiab] OR abortions[tiab] OR antenatal[tiab] OR babies[tiab] OR baby[tiab] OR birth[tiab] OR births[tiab] OR breast feeding[tiab] OR breastfeeding[tiab] OR childbearing[tiab] OR deliveries[tiab] OR delivery[tiab] OR gestation[tiab] OR gestational[tiab] OR health facilities[tiab] OR health facility[tiab] OR hospital[tiab] OR hospitals[tiab] OR "maternal Fetal infection transmission"[tiab] OR "maternal-fetal infection transmission"[tiab] OR midwife[tiab] OR midwifery[tiab] OR midwifes[tiab] OR midwive[tiab] OR midwives[tiab] OR miscarriage[tiab] OR miscarriages[tiab] OR mother[tiab] OR "Mother to child transmission"[tiab] OR "Mother to child transmissions"[tiab] OR mothers[tiab] OR neonate[tiab] OR neonates[tiab] OR newborn[tiab] OR newborns[tiab] OR obstetrical[tiab] OR obstetric[tiab] OR perinatal[tiab] OR post partum[tiab] OR postnatal[tiab] OR postpartum[tiab] OR pregnancies[tiab] OR pregnancy[tiab] OR pregnant[tiab] OR prenatal[tiab] OR puerperia[tiab] OR puerperium[tiab] OR "vertical pathogen transmission"[tiab] OR vertical transmissions[tiab] OR vertical transmission[tiab] OR reproductive[tiab] OR maternity[tiab] OR maternities[tiab]))

***##Concept male involvement in TIAB***

AND (men's involvement[tiab] OR men's participation[tiab] OR men's support[tiab] OR men's engagement[tiab] OR "partner involvement"[tiab] OR "partner participation"[tiab] OR "partner support"[tiab] OR "partner engagement"[tiab] OR "partners involvement"[tiab] OR "partners support"[tiab] OR "partner's involvement"[tiab] OR "partner's participation"[tiab] OR "partner's support"[tiab] OR "spouses support"[tiab] OR "father involvement"[tiab] OR "father participation"[tiab] OR "father support"[tiab] OR "father engagement"[tiab] OR "fathers involvement"[tiab] OR "fathers participation"[tiab] OR "fathers support"[tiab] OR "father's involvement"[tiab] OR "father's participation"[tiab] OR "father's support"[tiab] OR "father's engagement"[tiab] OR "husband involvement"[tiab] OR "husband participation"[tiab] OR "husband support"[tiab] OR "husbands involvement"[tiab] OR "husbands participation"[tiab] OR "husbands support"[tiab] OR "husband's involvement"[tiab] OR "husband's participation"[tiab] OR "husband's support"[tiab] OR "male involvement"[tiab] OR "male participation"[tiab] OR "male support"[tiab] OR "male engagement"[tiab] OR "males involvement"[tiab] OR "spousal involvement"[tiab] OR "spousal participation"[tiab] OR "spousal support"[tiab]))

***##Humans only***

NOT ("animals"[MeSH Terms] NOT "humans"[MeSH Terms])
