## Additional File 2 for "A systematic review of the concept “male involvement in maternal health” by natural language processing and descriptive analysis"

**Screening**

**Included**

**Eligibility**

**Identification**

Records identified through database searching
(n = (5277 )

1.CINAHL:1219

2. EMBASE: 1565

3.SCOPUS:1410

4.Web of Science:2397

5.PUBMED:1945

Additional records identified through WHO Reproductive Health Library (n=7)
(n =	)

Records after duplicates removed
(n = 3975 )

Records screened
(n =3975)

Records excluded
(n =3406)

Full-text articles assessed for eligibility
(n = 569)

Full-text articles excluded, with reasons
No full text available (n=39)

Wrong study design (n=47)

Wrong publication type (n=54)

No data collection during pregnancy/childbirth (n=11)

No quantitative data collection (n=11)

No measurement of MI (n=60)

Published before 2000 (n=60)

Low Quality (n=5)

Studies included in quantitative synthesis (meta-analysis)
(n = 282)
