## Additional File 3 for "A systematic review of the concept “male involvement in maternal health” by natural language processing and descriptive analysis"

**Additional File 3: Results Fisher exact test**

|  | Involvement | | | | |
| --- | --- | --- | --- | --- | --- |
|  | ANC attendance | Financial support or transport | Presence delivery | HIV testing | Psychosocial scale measurement (>10 items) |
| Participation | 0.60 | 0.12 | 0.77 | 0.54 | 0.37 |
| Engagement | 0.41 | 0.16 | 0.62 | 0.61 | 0.16 |
| Attendance | 0.08 | 0.34 | 1 | 1 | 1 |
| Presence | 0.004 | 0.34 | 0.004 | 0.35 | 0.16 |
| Support | <0.001 | 0.013 | <0.001 | <0.001 | <0.001 |

*Table 5 Results of the Fisher exact tests for assessing a difference in the use of indicators between studies using the term involvement, participation, engagement, attendance, presence and support.*
